# Accuracy of self-reported HIV testing history and awareness of HIV-positive status among people living with HIV in four Sub-Saharan African countries

**DOI:** 10.1101/2020.09.16.20196105

**Authors:** Yiqing Xia, Rachael M Milwid, Arnaud Godin, Marie-Claude Boily, Leigh F Johnson, Kimberly Marsh, Jeffrey W Eaton, Mathieu Maheu-Giroux

## Abstract

**Background:** In many countries in Sub-Saharan Africa, self-reported HIV testing history and awareness of HIV-positive status from household surveys are used to estimate the percentage of people living with HIV (PLHIV) who know their HIV status. Despite widespread use, there is limited empirical information on the sensitivity of those self-reports, which can be affected by non-disclosure.

**Methods:** Bayesian latent class models were used to estimate the sensitivity of self-reported HIV testing history and awareness of HIV-positive status in four *Population-based HIV Impact Assessment* surveys in Eswatini, Malawi, Tanzania, and Zambia. Antiretroviral (ARV) metabolites biomarkers were used to identify persons on treatment who did not accurately report their status. For those without ARV biomarkers, the pooled estimate of non-disclosure among untreated persons was 1.48 higher than those on treatment.

**Results:** Among PLHIV, the sensitivity of self-reported HIV testing history ranged 96% to 99% across surveys. Sensitivity of self-reported awareness of HIV status varied from 91% to 97%. Non-disclosure was generally higher among men and those aged 15-24 years. Adjustments for imperfect sensitivity did not substantially influence estimates of of PLHIV ever tested (difference <4%) but the proportion of PLHIV aware of their HIV-positive status was higher than the unadjusted proportion (difference <8%).

**Conclusions:** Self-reported HIV testing histories in four Eastern and Southern African countries are generally robust although adjustment for non-disclosure increases estimated awareness of status. These findings can contribute to further refinements in methods for monitoring progress along the HIV testing and treatment cascade.

## Introduction

Monitoring the HIV treatment and care cascade is central to the *Joint United Nations Programme on HIV/AIDS’* (UNAIDS) objective of ending the AIDS epidemic as a public health risk by 2030 (1). Routine tracking of population-level progress towards the UNAIDS’ 2020 90-90-90 and 2030 95-95-95 diagnostic, treatment, and viral load suppression targets can guide public health initiatives and improve programmatic efficiencies (2). However, estimating progress towards the first pillar of the targets –the percentage of people living with HIV (PLHIV) who know their HIV status– is challenging. In sub-Saharan Africa (SSA), where 67% of the 38 million PLHIV were estimated to reside in 2019 (3), measures of awareness are typically constructed from data about self-reported HIV testing behaviour or reported directly from nationally representative household surveys (4-8).

Consideration of the potential for measurement bias is needed when interpreting self-reported survey data. Studies have shown that self-reporting about sensitive information, such as an individual’s HIV testing history and HIV status, could be affected by non-disclosure (6, 9, 10). For example, inconsistencies have been documented in Kenya and Malawi between an individual’s self-reported data and biomarkers for metabolites of antiretrovirals (ARVs) and viral load suppression (5, 10). While previous studies have sought to validate the accuracy of self-reported HIV status (10-12), analyzing recent data on both non-disclosure of self-reported HIV testing history and HIV status among PLHIV is key to improving the validity of these estimates.

Surveys that collect both self-reported information and ARV biomarkers can be used to assess the accuracy of self-reported HIV testing histories and HIV awareness status. In this study, Bayesian latent class models are used to estimate the sensitivity of self-reported HIV testing history and awareness of HIV status among PLHIV based on the presence of detectable ARVs (13).

## Methods

### Study population

The *Population-based HIV Impact Assessment* (PHIA) surveys are nationally representative multistage household-based surveys designed to provide population-level information on the burden of HIV disease and to document the progress of HIV programs (14-17). All four PHIA surveys with available microdata on PLHIV aged 15+ years of age were included in our analysis: *Swaziland (Eswatini) HIV Incidence Measurement Survey 2* (2015-2016), *Malawi PHIA* (2015-2016), *Tanzania HIV Impact Survey* (2016-2017), and *Zambia PHIA* (2016).

### Self-reports and antiretroviral (ARV) status

Participants who reported having ever received the results of any HIV test were classified as *ever tested and received results* (hereafter referred to as *“ever tested”*; *see Table S1)*. Participants who reported having received a positive test result after any HIV test, were classified as *aware of HIV-positive status*. The specific laboratory algorithms used to detect ARVs varied across surveys, although all were analyzed in the same laboratory, and included drugs in the nationally recommended first- and second-line regimens: efavirenz, lopinavir, and nevirapine (14-17).

### Bayesian latent class models

Bayesian latent class models (18) were used to quantify the sensitivity of both self-reported HIV testing history and HIV status awareness among PLHIV. Cross-tabulations of self-reports with ARV biomarkers provide empirical information on their sensitivity among those with detectable ARVs (Figure 1A).

**Figure 1.**
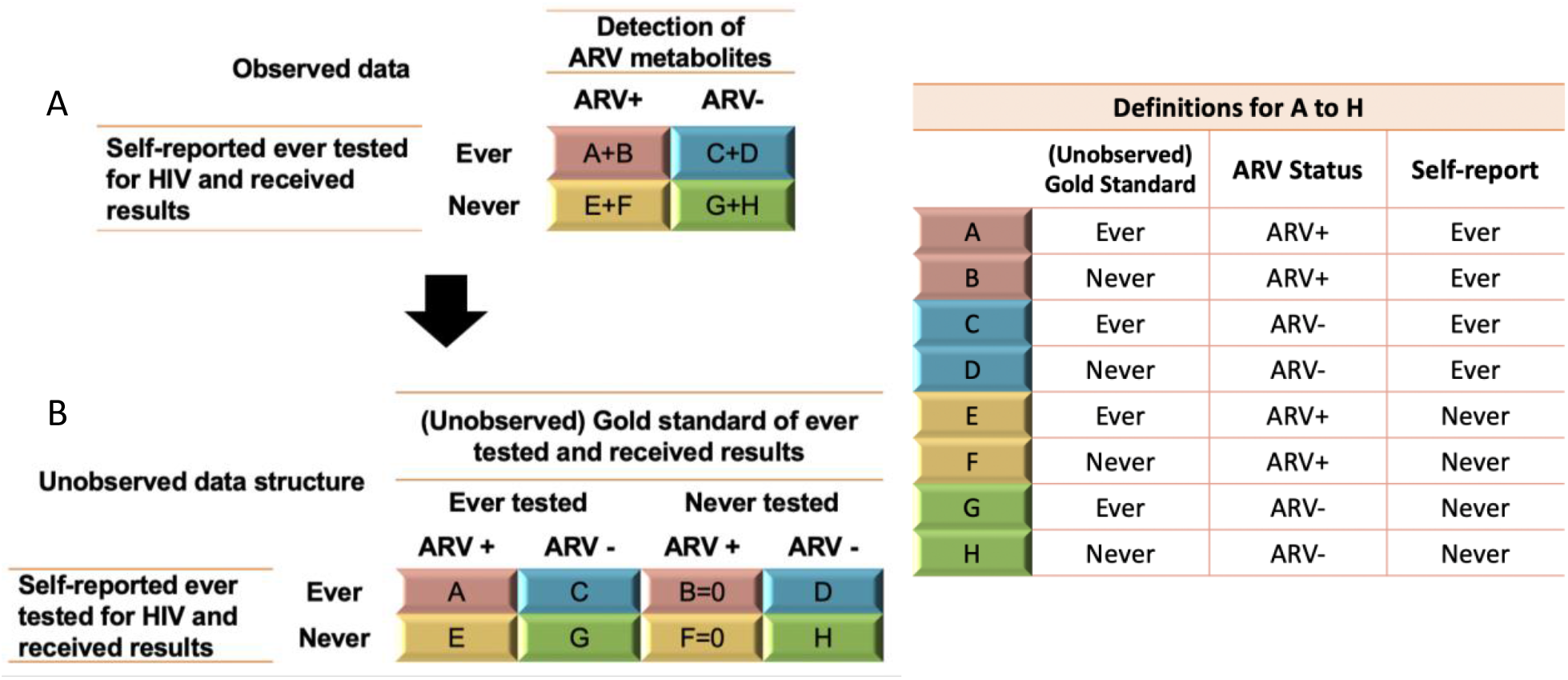
Observed and unobserved data structure of self-reported *ever tested and received results*, and antiretroviral (ARV) metabolites status. Definitions for cells A to H are listed in the table. As it was assumed that participants with detectable ARV must have been tested, cells B and F are *de facto* equal to zero. The same technique was applied to the self-reported awareness of HIV status and was applied to all four Sub-Saharan African countries.

With regard to participants with detectable ARVs, we assumed that: (1) they had been tested for HIV, received their results, and were aware of their status; and (2) there were no false positives in the detection of ARV metabolites, self-reported HIV testing history, or awareness of HIV status.

As ARV metabolite data only provide information about the sensitivity of self-reports among participants on ARVs, the ratio of non-disclosure for PLHIV without detectable ARVs versus those with detectable ARVs was given a log-normal prior distribution with a mean of *log(1*.*48)* (standard error: 4) to estimate the sensitivity of self-reports for people without detectable ARVs. This prior was elicited by reviewing available studies an meta-analyzing the evidence. The pooling of two studies conducted in rural Mozambique and Malawi (19, 20) suggests that people not receiving ARVs are 1.48 more likely to not disclose their diagnosis. Additional analyses were conducted to investigate the influence of this prior on our results. Equations and prior distributions are presented in the *Supplementary Materials (Table S2 and Text S1)*.

Given known biases in self-reported estimates of HIV status awareness, analysts often manually reclassify individuals not aware of their status but with detectable ARVs –as in published PHIA reports. Most surveys, however, do not collect ARV biomarkers and only rely on self-reported information. To examine the impact of this partial adjustment, we compared the unadjusted, ARV-reclassified (as in PHIA reports), and Bayesian-adjusted estimates of PLHIV aware.

Models were run separately for each country and for subgroup analyses (i.e. age, sex, urban/rural and socio-economic status). Bayesian hierarchical models using Markov Chain Monte Carlo (MCMC), implemented through the JAGS software (21) and the *rjags* packages, were used to approximate the posterior densities (22, 23).

### Ethics

Ethics approval for secondary data analyses was obtained from McGill University’s Faculty of Medicine Institutional Review Board (A10-E72-17B).

## Results

Overall, 3,003 PLHIV from Eswatini, 2,227 PLHIV from Malawi, 1,831 PLHIV from Tanzania, and 2,467 PLHIV from Zambia were included in the analyses. In all countries, a high fraction of PLHIV reported having ever been tested and the proportion of PLHIV reporting being aware of their status ranged from 68.0% in Tanzania to 86.5% in Eswatini. The proportion of PLHIV with detectable ARV metabolites was highest in Eswatini (76.0%), followed by Malawi (68.1%), Zambia (61.5%), and Tanzania (53.9%).

### Sensitivity of self-report

#### Self-reported testing history

Among participants with detectable ARVs, the estimated sensitivity was highest in Eswatini at 99.5% (95% credible interval [95%Crl]: 99.2-99.8%), followed by Malawi (98.2%; 97.5-98.8%), Zambia (97.4%; 96.5-98.1%), and Tanzania (96.6%; 95.3-97.6%) (Figure 2). For people without ARV metabolites, the estimated sensitivity was 2.4% points (0.1-11.4%) lower than those with detectable ARVs in Tanzania.The differences were smaller elsewhere. Detailed values can be found in *Supplementary Table 3*.

**Figure 2.**
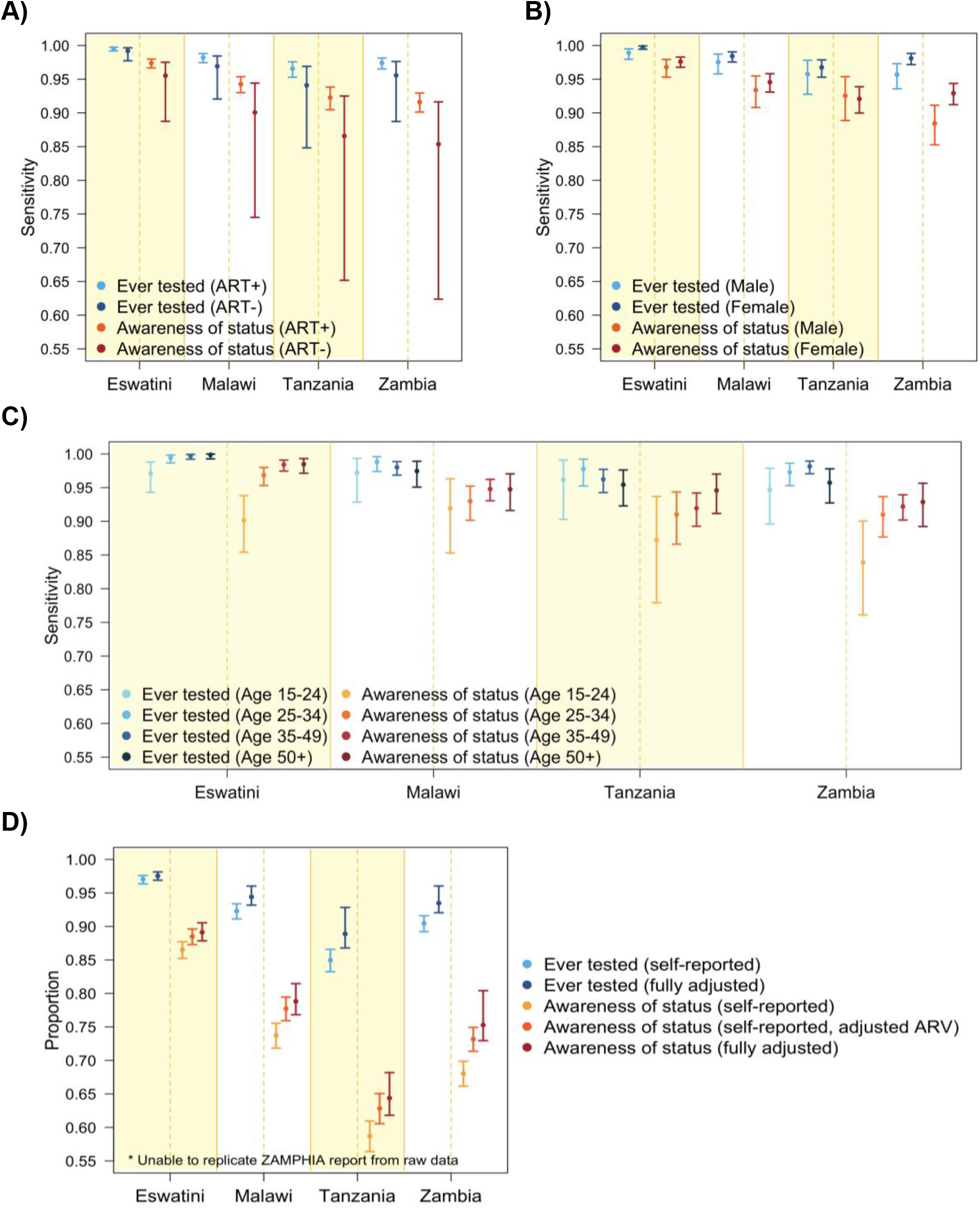
Marginal posterior medians and 95% credible intervals for selected outcomes. Sensitivity of self-reported ever tested and received results, and awareness of HIV-positive status among people living with HIV (PLHIV) by: (A) antiretroviral (ARV) metabolites status, (B) gender (ARV+), and (C) age groups (ARV+). Panel D portrays the overall proportion of PLHIV ever tested who received results and the awareness of HIV-positive status (self-reported vs. adjusted ARV metabolites status vs. fully adjusted).

#### Self-reported awareness of HIV status

The sensitivity of self-reported awareness of HIV-positive status among participants with ARV metabolites was 97.4% (96.7-98.0%) in Eswatini, 94.2% (93.0-95.4%) in Malawi, 92.3% (90.5-93.8%) in Tanzania, and 91.6% (90.1-92.9%) in Zambia (Figure 2A). The estimated differences in sensitivity between PLHIV with ARV metabolites and those without were 1.8% points (0.1-8.5%), 4.2% points (0.2-19.6%), 5.7% points (0.3-26.7%) and 6.2% points (0.3-29.0%) in Eswatini, Malawi, Tanzania, and Zambia respectively.

### Differences by gender, age, rural/urban, and socioeconomic status

Among participants with detectable ARVs, women had 0.9-2.4% points higher sensitivities of self-reported HIV testing history and HIV status awareness than men (Figure 2B). The estimated sensitivities were the lowest at age 15-24 years (94.7-97.2% for HIV testing history and 83.9-91.9% for HIV status awareness) in all of the countries (Figure 2C). Participants residing in urban and rural areas had similar sensitivities (*Figure S1A*) and variations by socio-economic status (SES) were also small (*Figure S1B and Figure S2*).

### Adjusted proportion of PLHIV ever tested and PLHIV aware of their status

Adjusting for imperfect sensitivity influenced the estimates of the self-reported proportion ever tested for HIV less (largest difference between the adjusted and the self-reports was 3.9% points in Tanzania) than the estimates of self-reported proportion of PLHIV aware of their status (largest difference was 7.2% points in Zambia) (Figure 2D). Results were less affected by the assumed non-disclosure ratio for PLHIV without detectable ARV metabolites when antiretroviral therapy (ART) coverage is high (*Figure S3*).

## Discussion

Self-reported information on HIV testing and diagnosis are primary data sources used to monitor trends in the HIV treatment and care continuum (2, 9). These same data have also been proposed to estimate cross-sectional HIV incidence (24). In this study, we leveraged ARV biomarkers from four household representative surveys in Eswatini, Malawi, Tanzania and Zambia to estimate the sensitivity of self-reported HIV testing history and awareness of HIV status among PLHIV. We found that self-reports of HIV testing history have a high sensitivity (>96%) among PLHIV with detectable ARVs across these settings. Self-reported awareness of HIV status had a marginally lower sensitivity (>91%) in these same countries.

Subgroup analyses revealed nonnegligible lower sensitivities of both self-reported HIV testing history and awareness of status among male PLHIV and those aged 15-24 years which could result from social desirability bias (25). Additionally, differences in survey instruments could result in higher sensitivities for women. For example, in the PHIA survey, women are asked about HIV testing up to 4 times (before pregnancy, during pregnancy, during labor, and at their last HIV test), while men were asked only once. As women were classified based on the positive response to any of the 4 questions, this increased the probability of women disclosing their true status.

In this study, we estimated the sensitivity of the self-reports alone but when ARV biomarkers are available, presentation of cascade results from the surveys usually adjust these self-reports by reclassifying PLHIV that do not disclose their status but for which ARV metabolites are detected as “aware”. We have found that this partial adjustment may be insufficient, especially if ART coverage is low in the surveyed population and the ratio of non-disclosure among those not on ART is high (26-29). To accurately estimate awareness of status, results must also be adjusted for non-disclosure among PLHIV with undetectable ARVs.

Our results need to be interpreted considering certain study limitations. First, only four PHIA surveys have publicly available micro-data, none of which are located in the West and Central African regions, where non-disclosure could be higher (30). The PHIA included here had some of the lowest levels of non-disclosure of these reviewed studies suggesting that other settings could have lower sensitivities. Second, our study design limited our assessment of the sensitivity of testing history to PLHIV, and findings should not be extrapolated to people not living with HIV. Third, it is not possible to empirically validate the sensitivity of self-reports among PLHIV without ARV metabolites. As such, we had to use information from two previous studies that used medical records to inform the non-disclosure ratio. Results could be sensitive to this non-disclosure ratio but the high ART coverage in the four countries mitigates this influence (*Figure S3*). Finally, the specificity of self-reports was assumed to be 100% which could lead to overestimating the proportion of PLHIV ever tested / aware of HIV-positive status. However, previous study has shown a high specificity of self-reported HIV testing results (11) implying that this assumption will likely have little impact on the outcomes.

Strengths of this study include the use of standardized survey and laboratory data (i.e. detection of ARV metabolites). Second, the Bayesian latent class models propagate uncertainty to our results by assuming prior distributions and generating posterior credible intervals. Finally, we examined sex, age, urban/rural, and SES differences in the sensitivity of self-reports.

In conclusion, self-reported HIV testing histories have high sensitivities in the four countries examined but self-reported awareness of HIV status are lower. Whenever available, ARV biomarkers data can be used to adjust self-reports but such adjustments may still underestimate diagnosis coverage, especially if ART coverage is low in that population. Future research should extend this work in other regions and populations.

## Data Availability

All data used in the manuscript is publicly accessible

## Acknowledgements

We acknowledge funding from the *Steinberg Fund for Interdisciplinary Global Health Research* (McGill University). MMG’s research program is funded through a *Canada Research Chair* (Tier 2) in *Population Health Modeling*. JWE acknowledges funding from the Bill and Melinda Gates Foundation and UNAIDS. MCB acknowledge funding from MRC Centre for Global Infectious Disease Analysis (MRC GIDA, MR/R015600/1). This award is jointly funded by the UK Medical Research Council (MRC) and the UK Department for International Development (DFID) under the MRC/DFID Concordat agreement and is also part of the EDCTP2 programme supported by the European Union. LJ acknowledges funding from UNAIDS.

## References

1. UNAIDS. 90 – 90 – 90: an ambitious treatment target to help end the AIDS epidemic. Geneva: Joint United Nationas Programme on HIV/AIDS (UNAIDS); 2014.

2. Rentsch CT, Georges Reniers, Richard Machemba, Emma Slaymaker, Milly Marston, Alison Wringe, et al. Non-disclosure of HIV testing history in population-based surveys: implications for estimating a UNAIDS 90-90-90 target. Global Health Action. 2018;11(1):1553470.

3. UNAIDS. UNAIDS data 2019.

4. Sarah Staveteig SW, Sara K. Head, Sarah E.K. Bradley, Erica Nybro. Demographic patterns of HIV testing uptake in sub-Saharan Africa: DHS comparative reports 30. Calverton: ICF Macro; 2013.

5. Kim AA, Mukui I, Young PW, Mirjahangir J, Mwanyumba S, Wamicwe J, et al. Undisclosed HIV infection and antiretroviral therapy use in the Kenya AIDS indicator survey 2012: relevance to national targets for HIV diagnosis and treatment. AIDS. 2016;30(17):2685–95.

6. Anand A, Shiraishi RW, Bunnell RE, Jacobs K, Solehdin N, Abdul-Quader AS, et al. Knowledge of HIV status, sexual risk behaviors and contraceptive need among people living with HIV in Kenya and Malawi. AIDS. 2009;23(12):1565–73.

7. Cherutich P, Kaiser R, Galbraith J, Williamson J, Shiraishi RW, Ngare C, et al. Lack of knowledge of HIV status a major barrier to HIV prevention, care and treatment efforts in Kenya: results from a nationally representative study. PLoS One. 2012;7(5):e36797.

8. Maheu-Giroux M, Marsh K, Doyle CM, Godin A, Laniece Delaunay C, Johnson LF, et al. National HIV testing and diagnosis coverage in sub-Saharan Africa: a new modeling tool for estimating the ‘first 90’ from program and survey data. AIDS. 2019;33 Suppl 3:S255–S69.

9. Organization WH. Consolidated strategic information guidelines for HIV in the health sector. Geneva: World Health Organization; 2015.

10. Fishel JD BB, Kishor S. Validity of data on self-reported HIV status and implications for measurement of ARV coverage in Malawi. DHS Working Paper No. 81. Calverton, Maryland, USA: ICF International; 2012.

11. Fisher DG, Reynolds GL, Jaffe A, Johnson ME. Reliability, sensitivity and specificity of self-report of HIV test results. AIDS Care. 2007;19(5):692–6.

12. Rohr JK, Xavier Gomez-Olive F, Rosenberg M, Manne-Goehler J, Geldsetzer P, Wagner RG, et al. Performance of self-reported HIV status in determining true HIV status among older adults in rural South Africa: a validation study. J Int AIDS Soc. 2017;20(1):21691.

13. Goncalves L, Subtil A, de Oliveira MR, do Rosario V, Lee PW, Shaio MF. Bayesian Latent Class Models in malaria diagnosis. PLoS One. 2012;7(7):e40633.

14. Malawi population-based HIV impact assessment (MPHIA) 2015-2016 data use manual supplement. New York, NY; December 2018.

15. Swaziland HIV incidence measurement survey 2 (SHIMS2) 2016-2017 data use manual supplement. New York, NY; April 2019.

16. Tanzania HIV impact survey (THIS) 2016-2017 data use manual supplement. New York, NY; December 2018.

17. Zambia population-based HIV impact assessment (ZAMPHIA) 2016-2017 data use manual supplement. New York, NY; February 2019.

18. Joseph L, Gyorkos TW, Coupal L. Bayesian estimation of disease prevalence and the parameters of diagnostic tests in the absence of a gold standard. Am J Epidemiol. 1995;141(3):263–72.

19. Fuente-Soro L, Lopez-Varela E, Augusto O, Sacoor C, Nhacolo A, Honwana N, et al. Monitoring progress towards the first UNAIDS target: understanding the impact of people living with HIV who re-test during HIV-testing campaigns in rural Mozambique. J Int AIDS Soc. 2018;21(4):e25095.

20. Chasimpha SJD, McLean EM, Dube A, McCormack V, Dos-Santos-Silva I, Glynn JR. Assessing the validity of and factors that influence accurate self-reporting of HIV status after testing: a population-based study. AIDS. 2020;34(6):931–41.

21. Plummer M, editor JAGS: A Program for Analysis of Bayesian Graphical Models Using Gibbs Sampling. Proceedings of the 3rd International Workshop on Distributed Statistical Computing (DSC 2003); 2003 March 20–22; Vienna, Austria.

22. Alan E. Gelfand AFMS. Sampling-based approaches to calculating marginal densities. Journal of the American Statistical Association. 1990;85(410):398–409.

23. Alan E. Gelfand SEH, Amy Racine-Poon, Adrian F. M. Smith. Illustration of Bayesian inference in normal data models using Gibbs sampling. Journal of the American Statistical Association. 1990;85(412):972–85.

24. Fellows IE, Shiraishi RW, Cherutich P, Achia T, Young PW, Kim AA. A new method for estimating HIV incidence from a single cross-sectional survey. PLoS One. 2020;15(8):e0237221.

25. Mooney AC, Campbell CK, Ratlhagana MJ, Grignon JS, Mazibuko S, Agnew E, et al. Beyond social desirability bias: investigating inconsistencies in self-reported HIV testing and treatment behaviors among HIV-positive adults in North West province, South Africa. AIDS Behav. 2018;22(7):2368–79.

26. Eswatini GotKo. Swaziland HIV incidence measurement survey 2 (SHIMS2) 2016-2017. Final report. Mbabane: Government of the Kingdom of Eswatini; April 2019.

27. Ministry of Health M. Malawi population-based HIV impact assessment (MPHIA) 2015-2016: Final report. Lilongwe: Ministry of Health; October 2018.

28. Tanzania Commission for AIDS (TACAIDS), (ZAC) ZAC. Tanzania HIV impact survey (THIS) 2016-2017: Final report. Dar es Salaam: Tanzania; December 2018.

29. Ministry of Health Z. Zambia population-based HIV impact assessment (ZAMPHIA) 2016: Final report. Lusaka: Ministry of Health; February 2019.

30. Eba PM. HIV-specific legislation in sub-Saharan Africa: A comprehensive human rights analysis. African Human Rights Law Journal. 2015;15:224–62.

